# GLOBAL PREVALENCE OF DEPRESSION IN CHRONIC KIDNEY DISEASE: A SYSTEMATIC REVIEW AND META-ANALYSIS

**DOI:** 10.1101/2023.09.03.23294994

**Authors:** Oluseyi Ademola Adejumo, Imuetinyan Rahsida Edeki, Oyedepo Dapo, Joshua Falade, Olawale Elijah Yisau, Olanrewaju Olumide Ige, Adedayo Oluwadamilola Adesida, Hansel Palencia, Ayman Moussa, Jibril Abdulmalik, Jean Jacques Noubiap, Udeme Ekpenyong Ekrikpo

## Abstract

**Background:** Chronic kidney disease (CKD) is commonly associated with psychosocial problems, especially depression, contributing to poor overall outcomes in CKD patients. Depression has not been given adequate priority in the management of CKD despite its significant adverse impact.

**Objectives:** This systematic review and meta-analysis determined the pooled prevalence of clinical depression in the global CKD population and sub-populations.

**Design:** A systematic search of PubMed, African Journals Online (AJOL), and EMBASE was undertaken to identify published articles with relevant data between 1989 and 2022. The pooled prevalence of clinical depression in the global CKD population was determined using random effects meta-analytic techniques.

**Participants:** Global adult and paediatric CKD population

**Results:** Eligible Sixty-one articles were included in this review, comprising 79,691 CKD patients from 26 countries. The participants’ mean age ranged from 11.0 to 76.3 years. Most (68.9%) of the studies had medium methodological quality. The overall pooled prevalence of depression was 27.6% (95% CI: 23.9–31.5%). Studies using Diagnostic Statistical Manual (DSM), International Classification of Disease, Mini-International Neuropsychiatric Interview and Structured Clinical Interview for DSM disorder returned a pooled prevalence of 31.7%, 39.6%, 23.3%, 25.1%, respectively; p=0.09. There was significant difference in the pooled prevalence of CKD across the various continents; p=0.01.The prevalence of depression was higher among patients on chronic haemodialysis compared to those in pre-dialysis (31.1% versus 18.9%; p=0.02), and among those on hemodialysis compared to peritoneal dialysis (31.9% versus 20.4%; p=0.03). There was no significant difference between adults and children (28.0% versus 15.9%, p=0.17).

**Conclusion:** Depression is a common disorder in patients with CKD. The findings of this study have brought to the fore the need for clinicians to make deliberate efforts to evaluate CKD patients for depression, especially those with advanced stages of the disease.

The study protocol was registered with PROSPERO (CRD42022382708)

**Strengths and Limitations:** The pooled prevalence of depression in this study correctly represents the magnitude of the depression in the CKD population because it included only studies where depression was diagnosed clinically.

This review involved the global population of both adult and paediatric patients with pre-dialysis and dialytic kidney disease.

Only a few studies in this review determined the prevalence of depression in the early stages of CKD.

Studies reporting depression prevalence among kidney transplant patients were not included in this review, and the number of studies in the paediatric population was limited.

## INTRODUCTION

Chronic kidney disease (CKD) is a common disease affecting about 700 million people globally.[1] It is a significant cause of mortality worldwide.[2, 3] The burden of CKD is higher in low- and middle-income countries compared to high-income countries.[3] It causes a huge economic, physical, and psychosocial burden for the patients.[4,5] Psychosocial problems such as depression and anxiety are more commonly encountered in CKD patients and their caregivers compared to the general population.[6,7] These problems adversely affect the quality of life and contribute to deterioration in renal function, progression to an advanced stage of disease, hospitalisation, and mortality in CKD patients.[8,9] Depression may contribute to poor compliance with adherence with medications, fluid, and dietary prescriptions of patients with CKD.[10,11] It also plays a pivotal role in the development of cardiovascular disease in CKD.[12] Psychosocial problems adversely affect the quality of life of CKD patients and contribute to overall poor outcomes of CKD.[6,13]

Despite the significant impact of psychosocial problems on the overall outcomes of CKD, they have not been given adequate priority in the management of CKD patients. Depression is not routinely screened for and managed in these patients, and mental health professionals were rarely mentioned in their multidisciplinary management.[14] Although there is inertia in the use of medications to manage psychosocial problems such as depression in CKD due to safety concerns by most clinicians, evidence supporting the safety of some medications and associated beneficial effects is now emerging.[15] In addition, non-pharmacological therapy, such as psychosocial interventions, has been established to improve the quality of life and reduce depression and anxiety in CKD patients.[16] Routine evaluation for common psychosocial problems, especially depression in this at-risk group, will aid in prompt diagnosis and allow those affected to be managed with consequent improvement in their overall outcomes.

This study assessed the prevalence of clinical depression in the global chronic kidney disease population. The findings of the study will provide helpful information on the burden of depression and also serve as empirical evidence to include its routine evaluation and treatment in the management of CKD, especially in climes where this is not a norm.

## METHODS

The Preferred Reporting Items for Systematic Review and Meta-Analysis (PRISMA) guideline [17] was used in reporting this study (Supplementary Table 1). This study was registered with PROSPERO with registration number CRD42022382708.

### Literature search

A systematic literature search was done on published articles involving CKD patients with clinically diagnosed depression. We performed a search on PubMed, African Journal Online, and Embase using terms related to depression in chronic kidney disease such as “chronic kidney disease”, “CKD”, “chronic renal failure”, “CRF”, “end-stage renal disease, “ESRD”, “end-stage kidney failure”, “ESKF”, “dialysis-dependent chronic kidney disease”, “renal failure” “non-dialysis dependent chronic kidney disease”, “pre-dialysis chronic kidney disease”, “chronic renal insufficiency”, “dialysis population” “depression”, “depressive symptom”, mental health disorder ”, psychiatric disorder’ in conjunction with all the names of continents were used in the search. The search strategy is detailed in Supplementary Tables 2A and 2B.

### Study selection

We included all identified studies reporting on clinically diagnosed depression in adult and paediatric non-dialysis and dialysis CKD populations across all the continents published between 1989 and 2023 in the literature search. We excluded articles where depression was diagnosed using screening tools. Two investigators (OAA and OOI) independently screened records for eligibility based on titles and abstracts. Full texts of articles deemed potentially eligible were retrieved and screened by the same investigators (OAA and OOI) for final inclusion. All conflicts were resolved by a third investigator (OEY).

### Data extraction and management

The following variables were extracted from selected studies: the last name of the first author, year of publication and country and continent the study was done, the sample size of the study, duration of the study, study design, mean age of the study participants, stage of CKD, type of renal replacement therapy, the proportion with depression, clinical depression diagnostic criteria used [Diagnostic Statistical Manual III (DSM III), DSM IV, DSM V, International Classification of Disease-9 (ICD-9), ICD-10, mini-international neuropsychiatric interview (MINI), structured clinical interview for DSM disorder (SCID)] and severity of depression. We categorised study location based on the World Health Organization regions [(African Region, AFR), (Region of the Americas, AMR), (South East Asian Region, SEAR), (European Region, EUR), (Eastern Mediterranean Region, EMR), and (Western Pacific Region, WPR)]

### Methodological quality

The Joanna-Briggs Institute Critical Appraisal Checklist for Studies Reporting Prevalence Data was used to assess the methodological quality of the constituent studies.[18] Studies scored 1 for each of the 9 questions with a “yes” response. Studies with scores 0 to 3 were regarded as poor quality, 4 to 6 as intermediate or medium quality, and 7 to 9 as high quality.

### Statistical analysis

Stata 17.0 (Stata Corp., 2021. Stata Statistical Software: Release 17, College Station, TX) was used for statistical analysis. The pooled prevalence of depression in the global CKD population was determined using meta-analytic techniques. The study-specific estimates derived from the DerSimonian-Laird random effects model[19] were pooled to estimate the prevalence of depression in this population. To minimise the effect of extreme values, the Freeman-Tukey double arcsine transformation[20] was used to stabilise the individual study variances before using the random effects model to obtain the pooled estimates. Publication bias was assessed using the Egger test.[21] We also undertook a subgroup analysis of pooled prevalence by continent, HD versus PD population, pre-dialysis versus dialysis CKD population, paediatric versus adult CKD population, among studies that used different clinical diagnostic tools such as DSM III, DSM IV, DSM-V, ICD-9, ICD-10, MINI, and SCID. Subgroup analysis was performed using the Q-test based on ANOVA. The I^2^ statistic was used to determine the between-study heterogeneity.

### Patient and public involvement

Patients and/or the public were not involved in the design, conduct, reporting, or dissemination plans of this research.

## RESULTS

### Study selection and characteristics

The systematic literature search initially identified 9,240 articles, of which 133 were selected for full-text review after duplicate removal and title and abstract screening. Finally, 61 articles [22–82] were eligible and included in this systematic review (Figure 1), with publication years ranging from 1989 to 2023. Included studies reported on 79,691 CKD and kidney failure patients from 26 countries. There were 249

**Figure 1:**
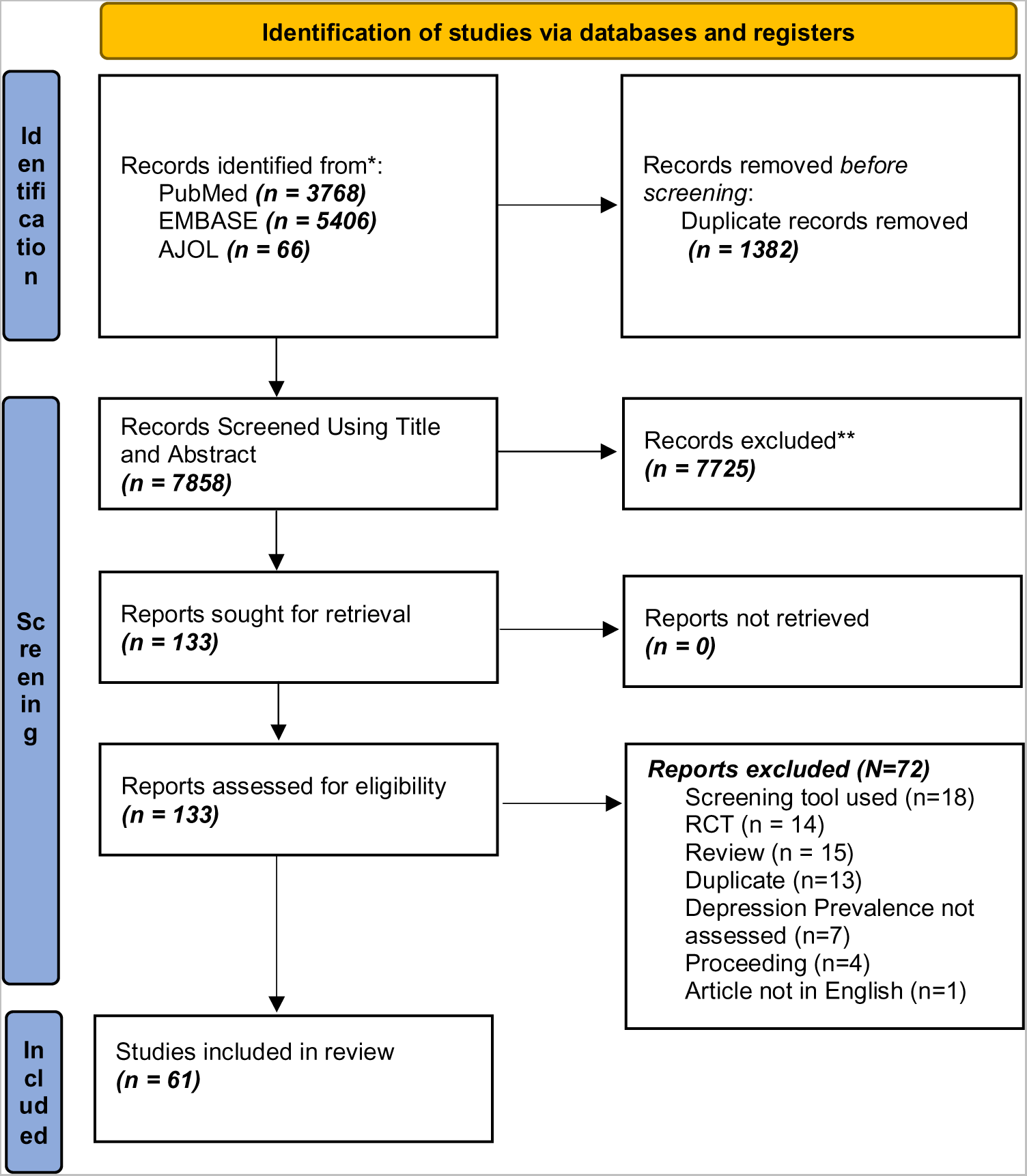
PRISMA flow diagram – Identification and Screening of Articles.

Participants (3 studies) from AFR [30,58,70]; 4,295 (14 studies) from EUR [31,33,34,36,39,43,56,61,66,68,75,78,80,82]; 3,962 (17 studies) from AMR [22,23,24,25,26,29,32,35,38,45,53,55,62,65,67,69,77]; 679 (7 studies) from EMR [40,49,51,60,64,74,81]; 846 (7 studies) from SEAR [28,47,50,59,71,73,79], and 69,660 (13 studies) from the WPR. [27,37,41,42,44,46,48,52,54,57,63,72,76] Figure 2 shows the geographical distribution of the continents of the included studies.

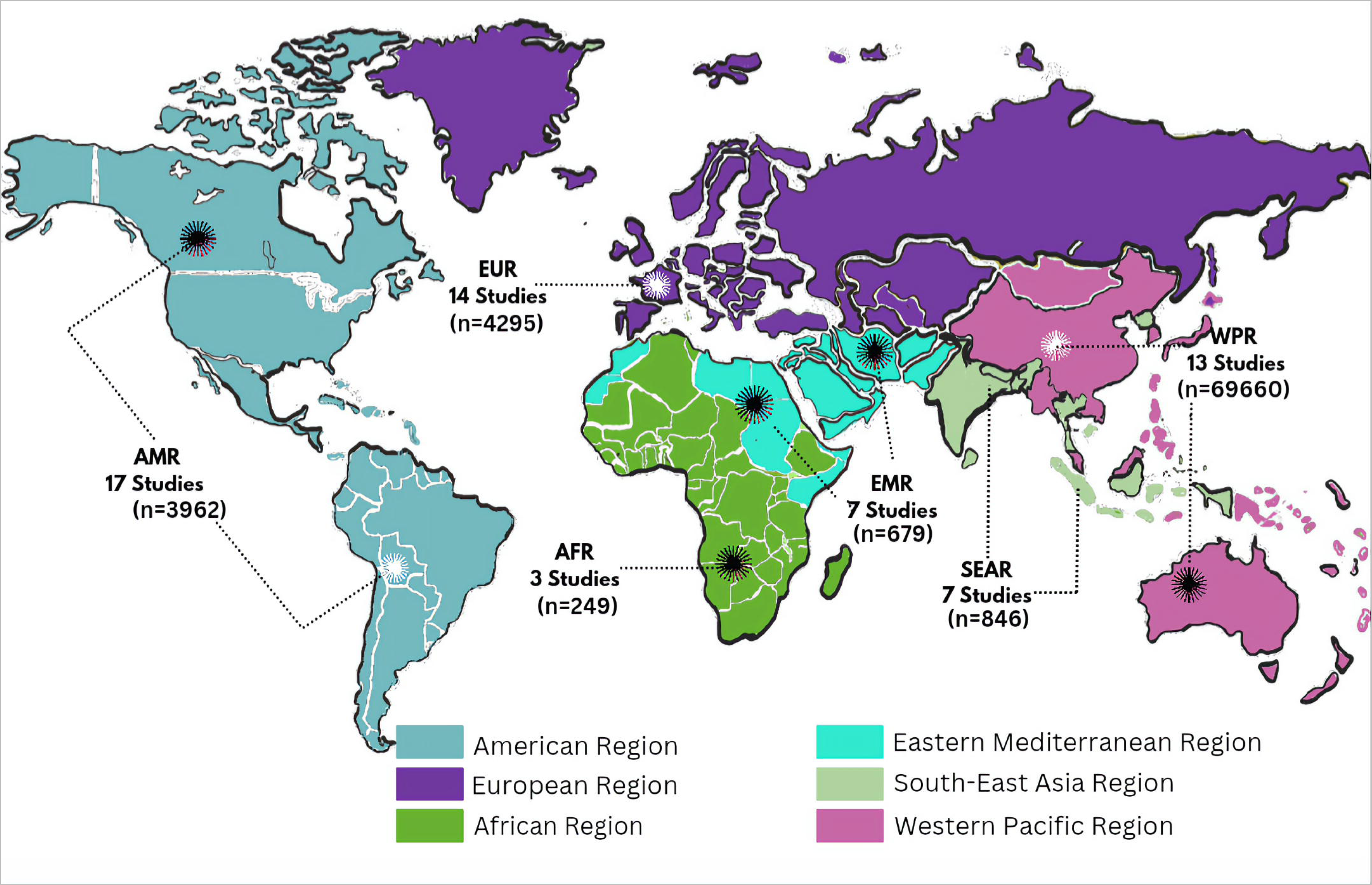

The clinical diagnostic criteria included those of the DSM (20 studies, 2,028 participants) [23,26,27,28,32,34,35,42,46,49,56,60,66,67,70,72,73,74,76,78] ICD (5 studies, 69,954 participants) [25,37,47,71,79] SCID (15 studies, 1,435 participants)[22,33,39,40,43,48,51,53,54,56,57,59,61,65,82], and MINI (21 studies, 6,274 participants). [24,29,30,31,36,38,41,44,45,50,52,55,58,62,64,68,69,75,77,80,81] The sample size of the component studies ranged from 20 [70] to 67,866 [37] patients, with the proportion of women ranging from 0% [25] to 68% [74]. The mean age of the participants ranged from 11.0 years [60] to 76.3 years [77].

Most of the studies had medium methodological quality (68.9%, n = 42) (Table 1); 14 studies (22.9%) were of high quality, including three studies from the Americas [23,25,77], one study from the African region[30], two studies from the Western Pacific region[43,76], three studies from the Eastern Mediterranean region [64,74,81], three from the European region [61,75,80] and two studies from South East Asia.[71,79] Table 1 summarises data extracted from the constituent articles. The majority (39, 63.9%) of the articles reported on severe or major depressive illness only; 5 (8.2%) reported on all spectrum of depression; 2 (3.3%) reported only on mild or moderate depression, while 15 (24.6%) did not state depression severity.

**Table 1:**
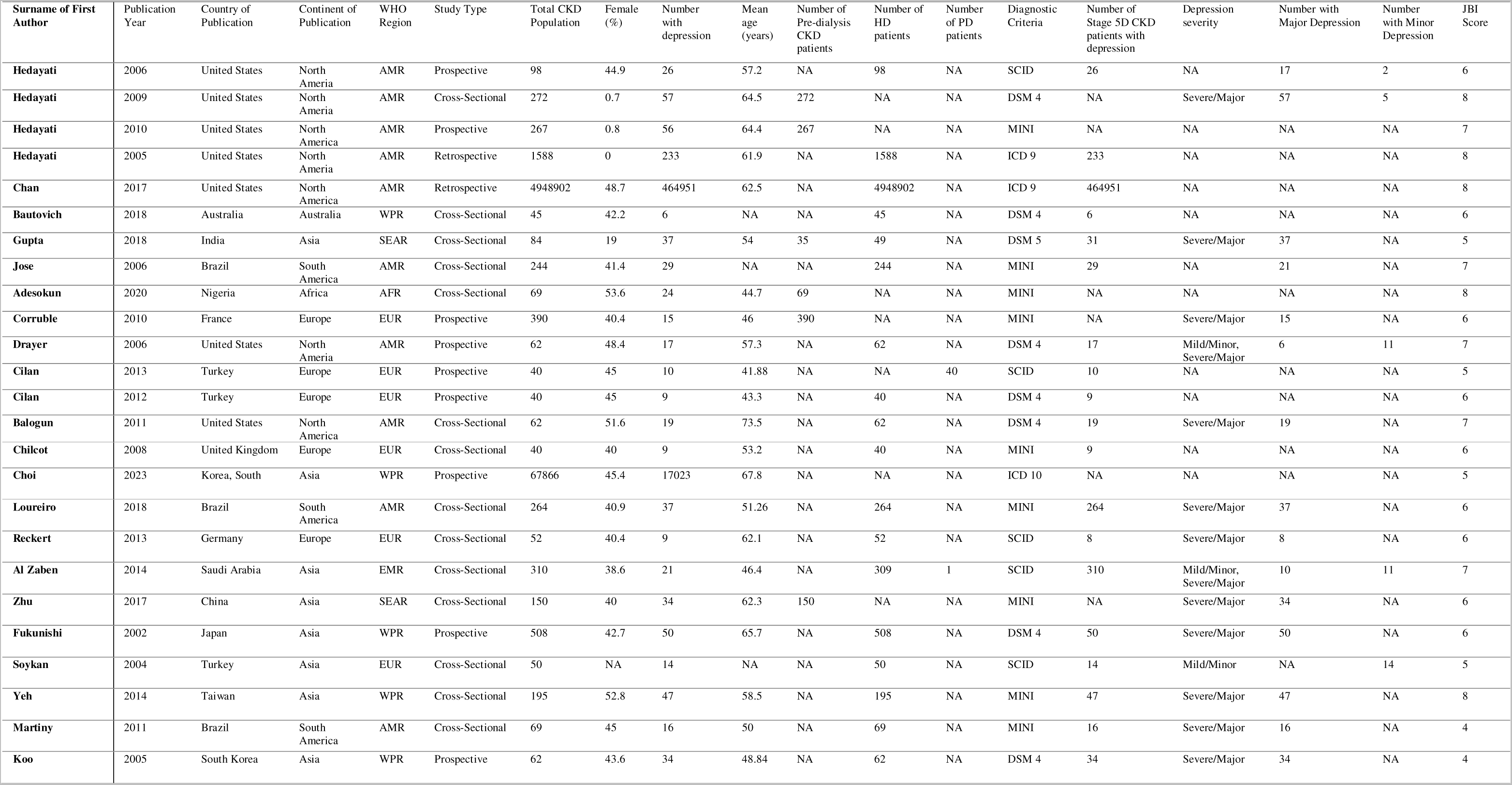
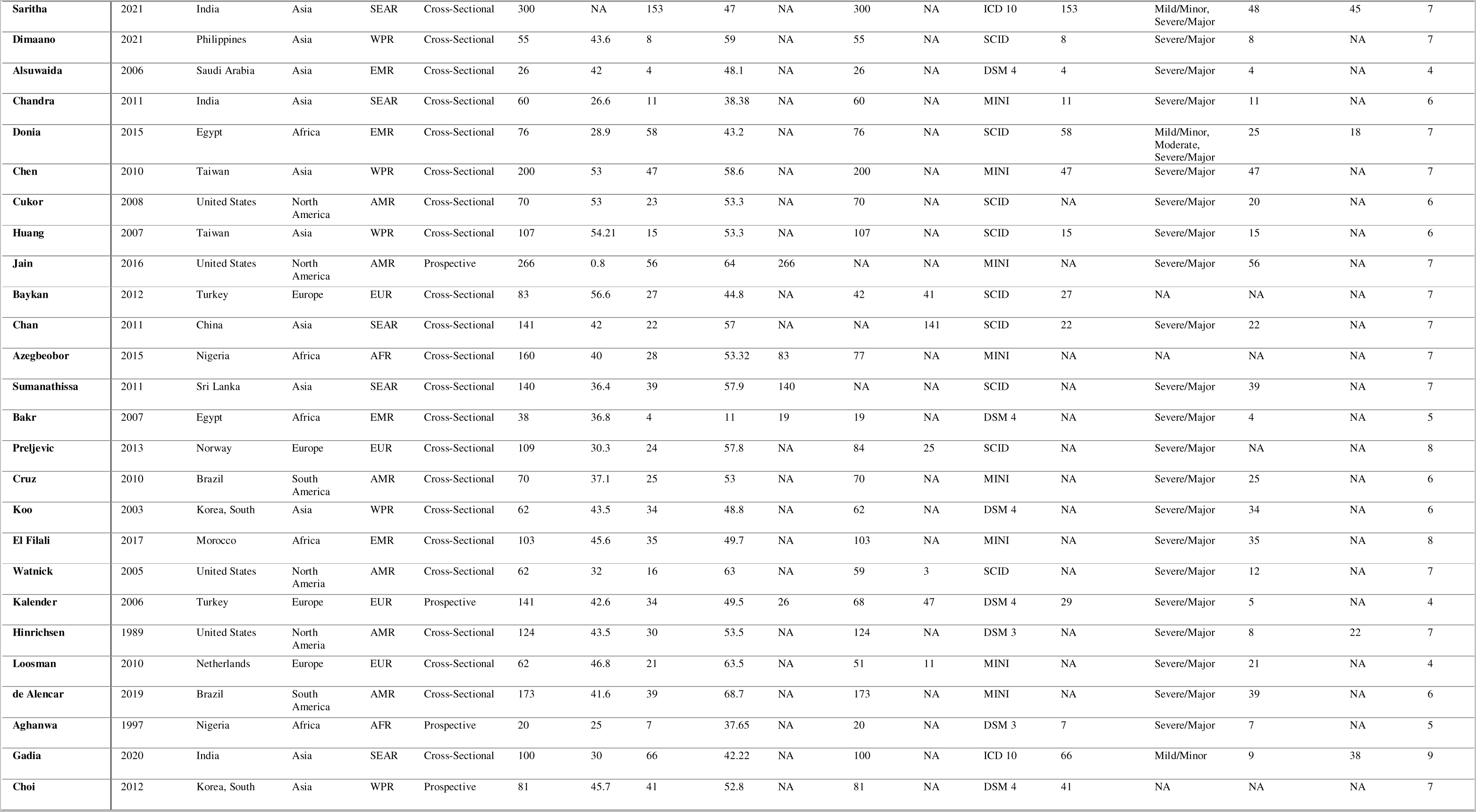
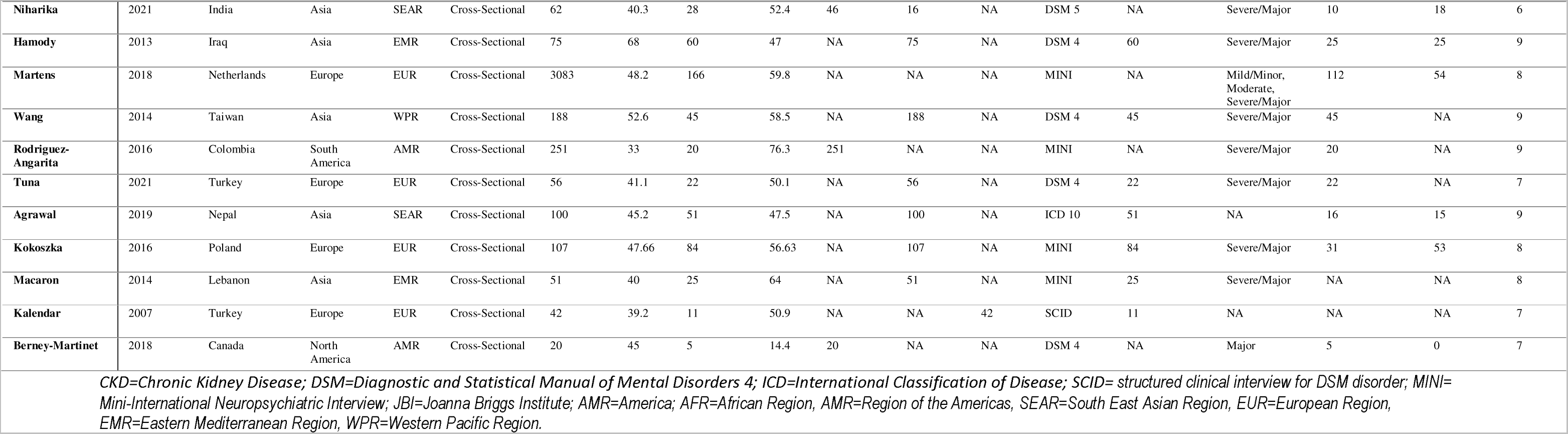
Summary of Extracted Data from Included Articles.

### Prevalence of depression

The overall pooled prevalence of depression was 27.6% [95% CI 23.9 – 31.5%), N = 61 studies, I^2^ = 97.3%, p<0.001 for heterogeneity] irrespective of the clinical depression diagnostic criteria used. Studies using DSM, ICD, MINI and SCID returned a pooled prevalence of [31.7% (23.4 – 40.6%), n = 20 studies, I^2^ = 93.7%, p<0.001], [39.6% (28.1-51.7%), n = 5 studies, I^2^ = 98.7%], [23.3% (16.5 – 30.8%), n = 21 studies, I^2^ = 97.1%], [25.1% (17.1 – 34.0%), n = 15 studies, I^2^ = 92.5%, p<0.001, respectively; p = 0.09 for difference across the different diagnostic criteria (Figure 3). The p-value for the Egger test was 0.83, suggesting no small study effects.

**Figure 3:**
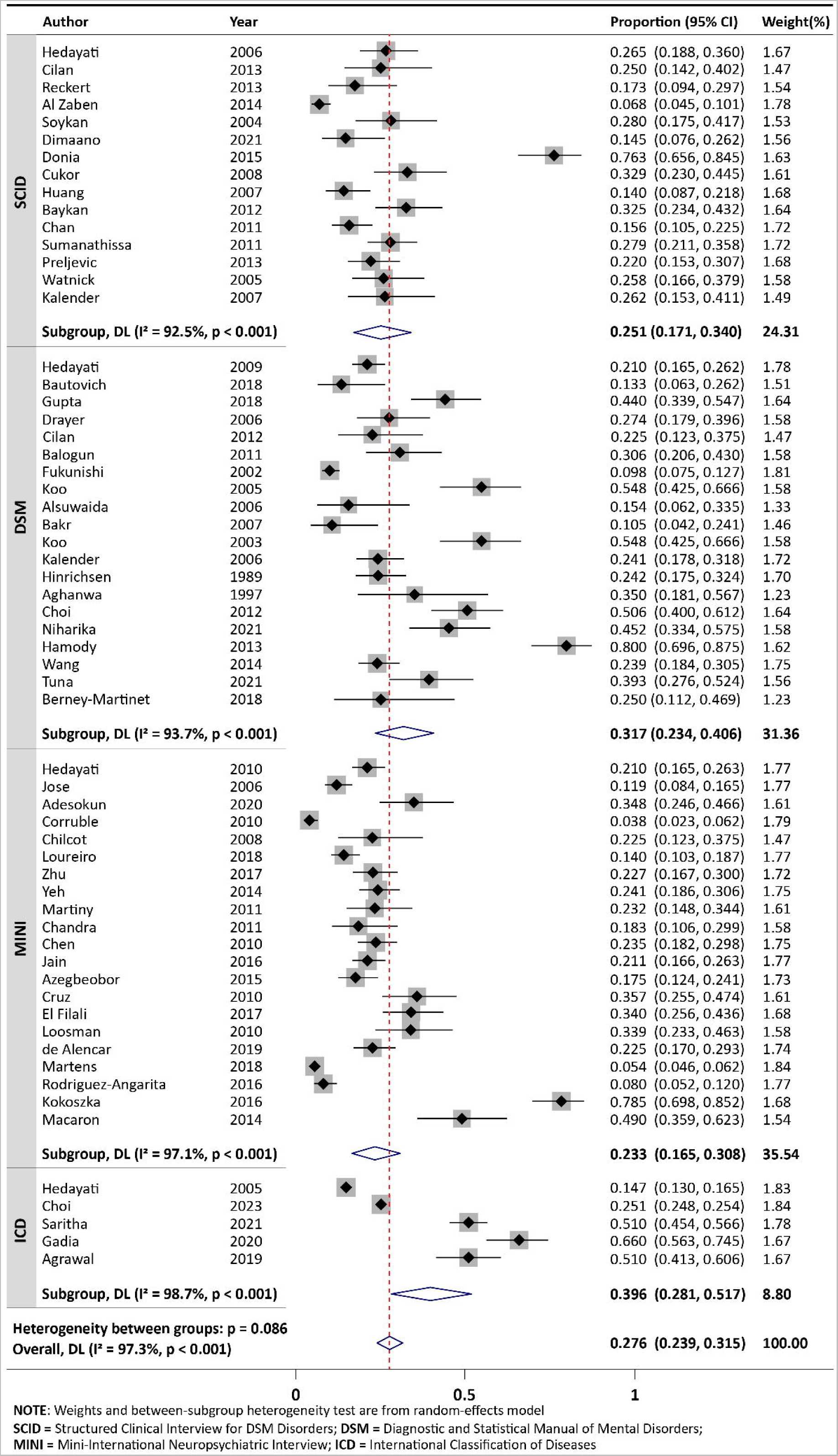
Forest plot showing the pooled prevalence of depression in CKD patients across diagnostic criteria.

The SEAR had the highest pooled prevalence of 43.2% (32.2 – 54.5%, I^2^ = 90.2%, p<0.001, n = 7 studies) compared to AMR 21.1% (17.6-24.7%, n=17 studies, I^2^=82.8%, p<0.001), WPR 25.0% (19.6-30.8%, n=13 studies, I^2^=93.3%), AFR 27.2% (14.2 – 42.5%, n = 3 studies, I^2^ = 78.4%, p=0.01), EUR 25.7% (14.8 – 38.3%, n = 14 studies, I^2^ = 97.5%, p<0.001); p=0.01 for difference across groups (Supplementary Figure 1).

There was a significantly higher pooled prevalence of clinical depression among the patients on chronic haemodialysis compared to pre-dialysis patients 31.1% (25.3-37.2%, n = 42 studies, I^2^ = 95.7%, p<0.001) versus 18.9% (11.9-27.1%, n = 10 studies, I^2^ = 93.8%, p<0.001); p=0.02 for difference across the groups (Supplementary Figure 2).

There was no significant difference in the pooled prevalence among adults compared with children (p = 0.17), Supplementary Figure 3. There was a higher prevalence of depression among the hemodialysis than the peritoneal dialysis population, 31.9% (26.0-38.1%, n=41 studies, I^2^ = 95.6%, p< 0.001) versus 20.4% (13.1-28.7%, n=3 studies, I^2^ = 41.7%, p=0.18); p=0.03 for difference between the groups, Supplementary Figure 4.

## DISCUSSION

This systematic review and meta-analysis of 61 studies determined the pooled prevalence of clinical depression among 79,691 CKD and kidney failure patients from 26 countries spread across the globe. The pooled prevalence of clinical depression was 27.6%. The prevalence varied with the geographical location of the CKD population, the stage of CKD, the age group of the CKD patient, the dialysis modality (haemodialysis versus peritoneal dialysis), and the clinical criterion used for the clinical diagnosis.

The pooled prevalence of depression in the CKD population in this study is higher than the prevalence of 8.4% and 6.9% reported in the general adult population in the United States and Norway, respectively.[83,84] Similarly, the prevalence of depression in the paediatric CKD population is higher than the 3.2% reported in the general children population in the United States.[85] These findings showed that depression is a common mental health problem in the CKD population compared to the general population. The prevalence of depression in this study is higher than 21.9% and 16.3% reported among patients with epilepsy and cancer, respectively.[86,87] This showed that the magnitude of depression in CKD is higher than in some other chronic illnesses.

The pooled prevalence of depression in this study is higher than 21.4% and 22.8% reported in the pre-dialysis and dialysis CKD population, respectively, in a systematic review and meta-analysis by Palmer et al. a decade ago. [88] It is, however, lower than 62% reported in a systematic review and meta-analysis involving a CKD population in Iran.[89] The wide difference in the prevalence rates may be due to the method of assessment of depression and the stage of CKD patients that were studied. While the diagnosis of depression was made in the present study using standard clinical diagnostic criteria, the study in Iran used a validated depression screening tool for the diagnosis of depression. The Iranian study also involved only patients on maintenance HD, while the present study included both pre-dialysis and maintenance HD patients.

There was no significant difference between the pooled prevalence of depression in the adult and paediatric population. However, this finding should be interpreted with caution as only two studies involving the paediatric population were included in this analysis. In addition, the prevalence of depression from these studies could have been underestimated as depression may have been unrecognised in children who are largely unable to adequately express themselves. Also, the presentation of depression in children may not follow the typically known presentation as commonly seen in the adult population. This underscores the need for physicians to take these peculiarities into consideration in the management of paediatric CKD population.

The pooled prevalence of clinical depression across the various continents ranges between 21.1-43.2%, with SEAR having the highest while the AMR had the lowest. The AFR had the second-highest pooled prevalence of depression (27.2%). There was a significant difference in the pooled prevalence of depression in CKD across the various continents. The difference in the severity of CKD, age and socio-economic status of the CKD population, available psychosocial support, governmental support, type of available health care financing, level of health care sophistication, type of maintenance HD and race may be partly responsible for the significant variations in prevalence of depression across the continents. The African continent had the least number of studies, while AMR had the highest number of studies. This underscores the need for more studies on depression in the African continent in order to ascertain the magnitude of the problem in the region.

In this systematic review and meta-analysis, the more commonly used clinical criteria were MINI and DSM, while ICD was the least used. Although the prevalence of depression varied between 23.3% and 39.6% depending on the clinical criteria used for diagnosis, there was no statistically significant difference across the various diagnostic criteria. This may suggest that any of the clinical criteria could be used in the assessment of depression in CKD.

The use of clinical methods of assessment is more specific than the use of validated screening tools for diagnosis of depression in the CKD population. Depression screening tools have been reported to over-diagnose depression in CKD patients, especially those at advanced stages.[88] The study by Palmer et al[88] is instructive in this regard, as it showed that the prevalence of depression was higher in CKD population when depression screening tools were used compared to when clinical diagnostic criteria were deployed. This was also buttressed by Smith et al[90] in their study, which showed prevalence rates of depression in the same HD population as 47% and 5% using Beck’s depression index and DSM III, respectively. Also, in cases where depression screening tools are being used in the assessment of depression in CKD patients, patients with advanced CKD may have some uremic symptoms such as poor sleep, anorexia, fatigue, and lack of concentration, which may overlap and be regarded as somatic symptoms of depression.[15,91] Another limitation of the use of depression screening tools in the diagnosis of depression lies in the fact that a higher score is commonly used as cut-off in advanced CKD compared to the general population.[15,91] These cut-off values may vary in different studies using the same screening tool, introducing some major inconsistencies.

There was a significantly higher pooled prevalence of depression among ESRD patients on haemodialysis compared to pre-dialysis patients (31.1% versus 18.9%) in this study. The demands of dialysis treatment on ESRD patients may partly account for this significant difference. Other factors such as dietary restriction, inflammation, hormonal changes, poor sleep quality, and reduced quality of life associated with dialysis treatment, may also contribute to a higher frequency of depression in HD patients compared to the pre-dialysis population.[16,15,91] A qualitative study by Avdal et al[92] also reported that ESRD patients experienced despair and decreased social support from relatives and caregivers after the commencement of PD or HD. This could effectively contribute to a higher prevalence of depression in them compared to the pre-dialysis population. This study also showed that about one in 5 pre-dialysis CKD patients had depression. This underscores the need to screen CKD patients, even in the early stages, for depression so that prompt treatment can be instituted. This may consequently improve their overall outcomes.

The pooled prevalence of depression was significantly higher in ESRD patients on maintenance HD compared with those on PD (31.9% versus 20.4%) in this study. There are conflicting reports from existing literature on the relationship between depression and mode of dialysis. The finding of this study is however supported by Martin et al.[93] but differs from some previous reports, which showed a significantly higher prevalence of depression in PD patients compared to those on HD.[94,95] Although some other studies reported a higher prevalence of depression in HD patients compared with PD patients, the difference was however not statistically significant.[96,97]

Depression has a significant adverse impact on the overall outcomes of CKD, hence it deserves adequate attention by being promptly diagnosed and managed by the clinician. Depression is associated with poor compliance with treatment follow-up; increased suicidal tendency, withdrawal from dialysis, medications dietary and fluid restrictions and malnutrition-inflammatory-atherosclerosis.[10,11,98,99,100] These may account for the increased hospitalisation and disease progression, reduced quality of life, and increased mortality in CKD patients who have depression.[6,8,9,13,101–103] Prompt diagnosis and management of depression may mitigate these adverse consequences and improve the overall outcome of these patients.

There is clinicians’ inertia in treating depression in CKD with antidepressants because of uncertainty about the pharmacokinetic properties of the medications and safety profile. This is due to the significant reduction in kidney function that is characteristic of the disease. This treatment inertia is supported by a report that showed that only about one-third of CKD patients diagnosed with depression received treatment.[104] Furthermore, most randomised control trials on efficacy and safety of antidepressants exclude CKD patients. Despite the above, there is some evidence, though limited, to support the effectiveness of antidepressants in reducing depression and improving QoL in CKD patients.[105] This present systematic review showed that clinical depression is common in CKD and lends credence to the need for the inclusion of CKD population in clinical trials on the efficacy of antidepressant treatment. Management of depression also involves non-pharmacological treatments. There are reports of some randomised control trials that showed the efficacy of non-pharmacological treatment of depression, such as cognitive-based therapy and relaxation techniques, in the treatment of depression in CKD.[16,106–108] Multidisciplinary team-based approach that includes mental health professionals will therefore be highly valuable in the management of CKD patients with depression.

The limitation of this study was that only a few studies in this review determined the prevalence of depression in early stages of CKD. Secondly, there was limited information on the prevalence of clinically diagnosed depression in the paediatric population, being reported in only 3 studies. The strength of this systematic review lies in the fact that the pooled prevalence of depression found in the CKD population is a fairly true representation of the magnitude of the disease. This is particularly true because the study included only articles that used clinical interviews which are more specific than screening tools to diagnose depression in CKD.

## CONCLUSION

This systematic review and meta-analysis showed that depression is a relatively common mental health disorder in the CKD population. It has brought to the fore the need for clinicians to make deliberate efforts to evaluate CKD patients, especially those with advanced stages of the disease for depression. We recommend a pragmatic approach of screening all CKD patients with the use of short and validated screening tests, especially in low- and middle-income countries with limited human resources for mental health care services. Those with depressive symptoms on screening should undergo clinical diagnosis before the commencement of depression management. Family support may immensely benefit CKD patients in low- and middle-income countries where the extended family system still exists. Secondly, considering the compelling evidence of the high prevalence of depression in patients with CKD, a form of universal intervention in collaboration with mental health professionals should be routinely provided. This may take the form of health talks while waiting for the clinic to commence, playing audio-visual materials, sharing informative fliers, and encouraging those struggling with their emotional well-being to indicate and be linked to mental health services. Lastly, the findings of this review have also given credence to the call to include CKD population in large, randomised control trials on the safety and efficacy of antidepressants because they are potential beneficiaries of the findings of such studies. While awaiting the recommendations from the ongoing RCTs, non-pharmacological treatment modalities such as CBT and relaxation techniques should be used in the treatment of those with depression.

## Authors’ contributions

OAA, UEE, IRE, FO and OD were involved in the conceptualisation of the study. All authors were involved in the literature search and review, methodology design and data collection. UEE was involved in data analysis. OAA, UEE, IRE, FO, OD, JF, HP, AM, JA, and JJN were involved in data interpretation. OAA and UEE were involved in writing the original draft. All authors were involved in the manuscript review and editing. All authors read and approved the final draft.

## Data Availability statement

The datasets generated during and/or analysed during the current study are available from the corresponding author upon reasonable request.

## Ethical Consideration

Ethical approval was not required. The study protocol was registered with PROSPERO (CRD42022382708).

## Data Availability

All data produced in the present work are contained in the manuscript.

## Acknowledgement

Nil

