## Supplementary Materials for "GLOBAL PREVALENCE OF DEPRESSION IN CHRONIC KIDNEY DISEASE: A SYSTEMATIC REVIEW AND META-ANALYSIS"

**Supplementary Fig. 1:** Forest plot showing the pooled prevalence across the WHO regions

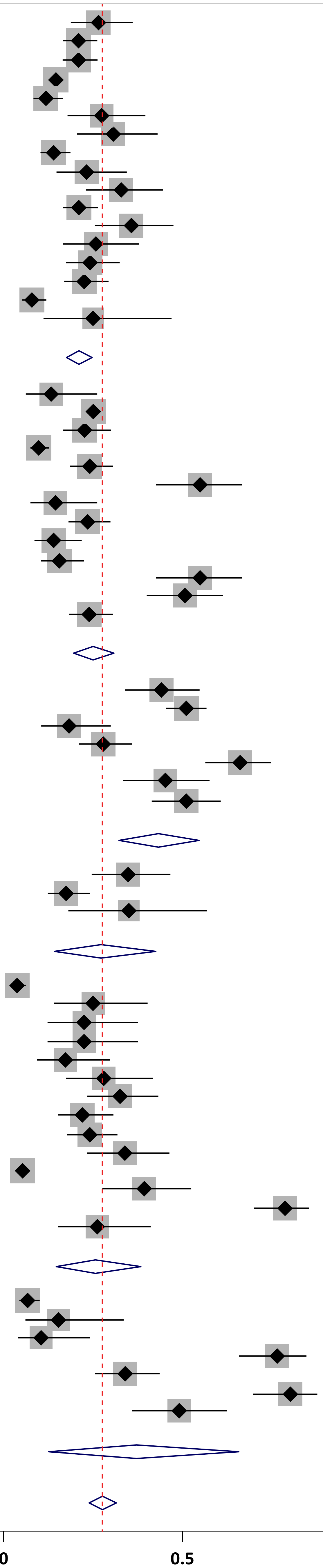


| Region | Author | Year | Proportion (95% CI) | Weight(%) |
|---|---|---|---|---|
| AMR | Hedayati | 2006 | 0.265 (0.188, 0.360) | 1.67 |
| | Hedayati | 2009 | 0.210 (0.165, 0.262) | 1.78 |
| | Hedayati | 2010 | 0.210 (0.165, 0.263) | 1.77 |
| | Hedayati | 2005 | 0.147 (0.130, 0.165) | 1.83 |
| | Jose | 2006 | 0.119 (0.084, 0.165) | 1.77 |
| | Drayer | 2006 | 0.274 (0.179, 0.396) | 1.58 |
| | Balogun | 2011 | 0.306 (0.206, 0.430) | 1.58 |
| | Loureiro | 2018 | 0.140 (0.103, 0.187) | 1.77 |
| | Martiny | 2011 | 0.232 (0.148, 0.344) | 1.61 |
| | Cukor | 2008 | 0.329 (0.230, 0.445) | 1.61 |
| | Jain | 2016 | 0.211 (0.166, 0.263) | 1.77 |
| | Cruz | 2010 | 0.357 (0.255, 0.474) | 1.61 |
| | Watnick | 2005 | 0.258 (0.166, 0.379) | 1.58 |
| | Hinrichsen | 1989 | 0.242 (0.175, 0.324) | 1.70 |
| | de Alencar | 2019 | 0.225 (0.170, 0.293) | 1.74 |
| | Rodriguez-Angarita | 2016 | 0.080 (0.052, 0.120) | 1.77 |
| | Berney-Martinet | 2018 | 0.250 (0.112, 0.469) | 1.23 |
| | **Subgroup, DL (I² = 82.8%, p < 0.001)** | | **0.211 (0.176, 0.247)** | **28.38** |
| WPR | Bautovich | 2018 | 0.133 (0.063, 0.262) | 1.51 |
| | Choi | 2023 | 0.251 (0.248, 0.254) | 1.84 |
| | Zhu | 2017 | 0.227 (0.167, 0.300) | 1.72 |
| | Fukunishi | 2002 | 0.098 (0.075, 0.127) | 1.81 |
| | Yeh | 2014 | 0.241 (0.186, 0.306) | 1.75 |
| | Koo | 2005 | 0.548 (0.425, 0.666) | 1.58 |
| | Dimaano | 2021 | 0.145 (0.076, 0.262) | 1.56 |
| | Chen | 2010 | 0.235 (0.182, 0.298) | 1.75 |
| | Huang | 2007 | 0.140 (0.087, 0.218) | 1.68 |
| | Chan | 2011 | 0.156 (0.105, 0.225) | 1.72 |
| | Koo | 2003 | 0.548 (0.425, 0.666) | 1.58 |
| | Choi | 2012 | 0.506 (0.400, 0.612) | 1.64 |
| | Wang | 2014 | 0.239 (0.184, 0.305) | 1.75 |
| | **Subgroup, DL (I² = 93.3%, p < 0.001)** | | **0.250 (0.196, 0.308)** | **21.89** |
| SEAR | Gupta | 2018 | 0.440 (0.339, 0.547) | 1.64 |
| | Saritha | 2021 | 0.510 (0.454, 0.566) | 1.78 |
| | Chandra | 2011 | 0.183 (0.106, 0.299) | 1.58 |
| | Sumanathissa | 2011 | 0.279 (0.211, 0.358) | 1.72 |
| | Gadia | 2020 | 0.660 (0.563, 0.745) | 1.67 |
| | Niharika | 2021 | 0.452 (0.334, 0.575) | 1.58 |
| | Agrawal | 2019 | 0.510 (0.413, 0.606) | 1.67 |
| | **Subgroup, DL (I² = 90.2%, p < 0.001)** | | **0.432 (0.322, 0.545)** | **11.65** |
| AFR | Adesokun | 2020 | 0.348 (0.246, 0.466) | 1.61 |
| | Azegbeobor | 2015 | 0.175 (0.124, 0.241) | 1.73 |
| | Aghanwa | 1997 | 0.350 (0.181, 0.567) | 1.23 |
| | **Subgroup, DL (I² = 78.4%, p = 0.010)** | | **0.272 (0.142, 0.425)** | **4.57** |
| EUR | Corruble | 2010 | 0.038 (0.023, 0.062) | 1.79 |
| | Cilan | 2013 | 0.250 (0.142, 0.402) | 1.47 |
| | Cilan | 2012 | 0.225 (0.123, 0.375) | 1.47 |
| | Chilcot | 2008 | 0.225 (0.123, 0.375) | 1.47 |
| | Reckert | 2013 | 0.173 (0.094, 0.297) | 1.54 |
| | Soykan | 2004 | 0.280 (0.175, 0.417) | 1.53 |
| | Baykan | 2012 | 0.325 (0.234, 0.432) | 1.64 |
| | Preljevic | 2013 | 0.220 (0.153, 0.307) | 1.68 |
| | Kalender | 2006 | 0.241 (0.178, 0.318) | 1.72 |
| | Loosman | 2010 | 0.339 (0.233, 0.463) | 1.58 |
| | Martens | 2018 | 0.054 (0.046, 0.062) | 1.84 |
| | Tuna | 2021 | 0.393 (0.276, 0.524) | 1.56 |
| | Kokoszka | 2016 | 0.785 (0.698, 0.852) | 1.68 |
| | Kalender | 2007 | 0.262 (0.153, 0.411) | 1.49 |
| | **Subgroup, DL (I² = 97.5%, p < 0.001)** | | **0.257 (0.148, 0.383)** | **22.48** |
| EMR | Al Zaben | 2014 | 0.068 (0.045, 0.101) | 1.78 |
| | Alsuwaida | 2006 | 0.154 (0.062, 0.335) | 1.33 |
| | Donia | 2015 | 0.763 (0.656, 0.845) | 1.63 |
| | Bakr | 2007 | 0.105 (0.042, 0.241) | 1.46 |
| | El Filali | 2017 | 0.340 (0.256, 0.436) | 1.68 |
| | Hamody | 2013 | 0.800 (0.696, 0.875) | 1.62 |
| | Macaron | 2014 | 0.490 (0.359, 0.623) | 1.54 |
| | **Subgroup, DL (I² = 97.9%, p < 0.001)** | | **0.371 (0.127, 0.656)** | **11.03** |
| | **Heterogeneity between groups: p = 0.006** | | | |
| | **Overall, DL (I² = 97.3%, p < 0.001)** | | **0.276 (0.239, 0.315)** | **100.00** |

**NOTE**: Weights and between-subgroup heterogeneity test are from random-effects model
**AMR**=Region of the Americas; **WPR**=West Pacific Region; **SEAR**=South-East Asian Region; **AFR**=African Region; **EUR**=European region; **EMR**=Eastern Mediterranean Region

**Supplementary Fig. 2:** Forest plot showing comparison of the pooled prevalence of depression in Pre-KRT versus KRT populations

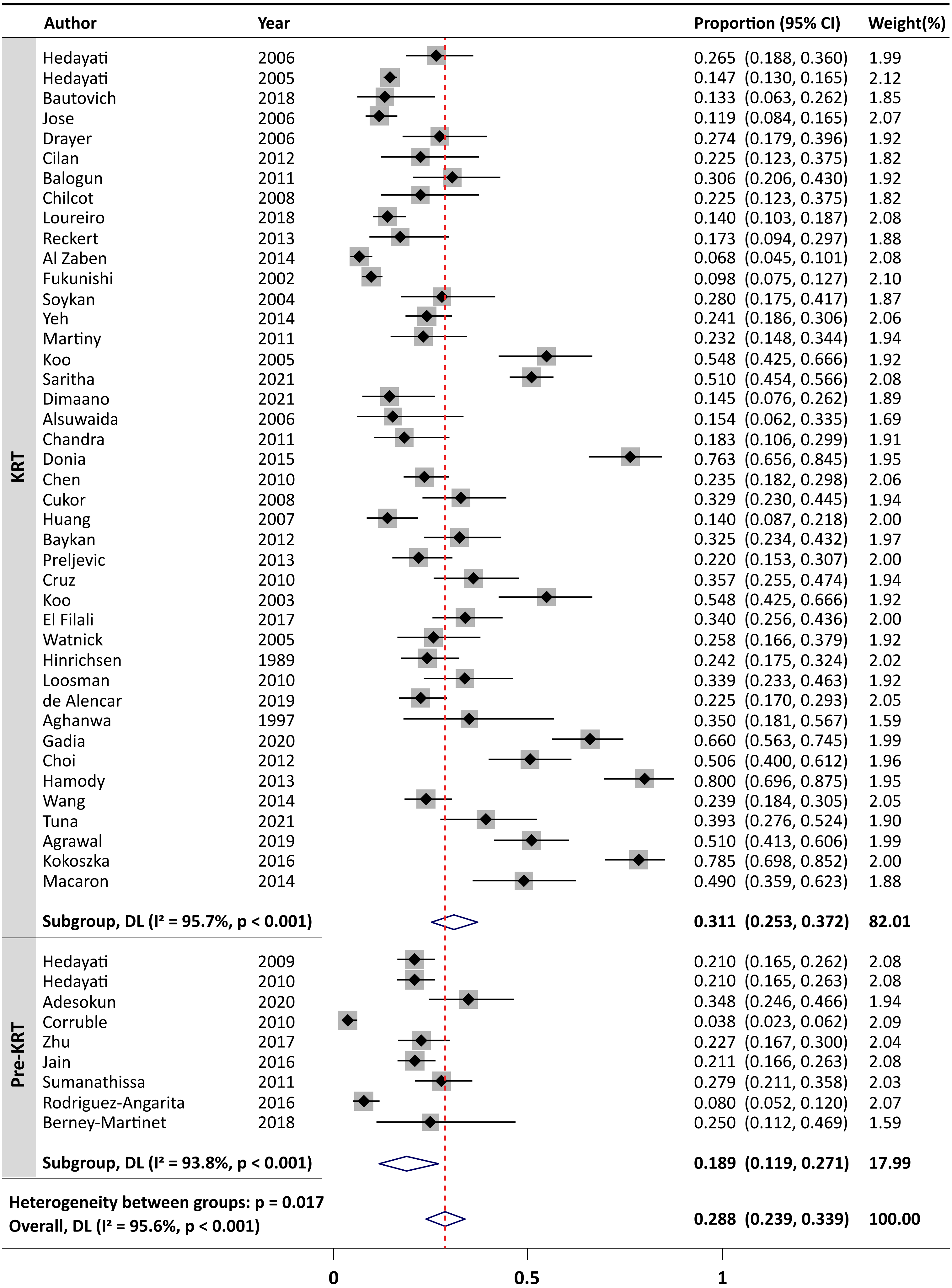


**NOTE**: Weights and between-subgroup heterogeneity test are from random-effects model

**KRT** = Kidney Replacement Therapy

**Supplementary Fig. 3:** Forest plot comparing the pooled prevalence of depression in adult versus paediatric CKD patients

| | Author | Year | Proportion (95% CI) | Weight(%) |
|---|---|---|---|---|
| Adult | Hedayati | 2006 | 0.265 (0.188, 0.360) | 1.67 |
| | Hedayati | 2009 | 0.210 (0.165, 0.262) | 1.78 |
| | Hedayati | 2010 | 0.210 (0.165, 0.263) | 1.77 |
| | Hedayati | 2005 | 0.147 (0.130, 0.165) | 1.83 |
| | Bautovich | 2018 | 0.133 (0.063, 0.262) | 1.51 |
| | Gupta | 2018 | 0.440 (0.339, 0.547) | 1.64 |
| | Jose | 2006 | 0.119 (0.084, 0.165) | 1.77 |
| | Adesokun | 2020 | 0.348 (0.246, 0.466) | 1.61 |
| | Corruble | 2010 | 0.038 (0.023, 0.062) | 1.79 |
| | Drayer | 2006 | 0.274 (0.179, 0.396) | 1.58 |
| | Cilan | 2013 | 0.250 (0.142, 0.402) | 1.47 |
| | Cilan | 2012 | 0.225 (0.123, 0.375) | 1.47 |
| | Balogun | 2011 | 0.306 (0.206, 0.430) | 1.58 |
| | Chilcot | 2008 | 0.225 (0.123, 0.375) | 1.47 |
| | Choi | 2023 | 0.251 (0.248, 0.254) | 1.84 |
| | Loureiro | 2018 | 0.140 (0.103, 0.187) | 1.77 |
| | Reckert | 2013 | 0.173 (0.094, 0.297) | 1.54 |
| | Al Zaben | 2014 | 0.068 (0.045, 0.101) | 1.78 |
| | Zhu | 2017 | 0.227 (0.167, 0.300) | 1.72 |
| | Fukunishi | 2002 | 0.098 (0.075, 0.127) | 1.81 |
| | Soykan | 2004 | 0.280 (0.175, 0.417) | 1.53 |
| | Yeh | 2014 | 0.241 (0.186, 0.306) | 1.75 |
| | Martiny | 2011 | 0.232 (0.148, 0.344) | 1.61 |
| | Koo | 2005 | 0.548 (0.425, 0.666) | 1.58 |
| | Saritha | 2021 | 0.510 (0.454, 0.566) | 1.78 |
| | Dimaano | 2021 | 0.145 (0.076, 0.262) | 1.56 |
| | Alsuwaida | 2006 | 0.154 (0.062, 0.335) | 1.33 |
| | Chandra | 2011 | 0.183 (0.106, 0.299) | 1.58 |
| | Donia | 2015 | 0.763 (0.656, 0.845) | 1.63 |
| | Chen | 2010 | 0.235 (0.182, 0.298) | 1.75 |
| | Cukor | 2008 | 0.329 (0.230, 0.445) | 1.61 |
| | Huang | 2007 | 0.140 (0.087, 0.218) | 1.68 |
| | Jain | 2016 | 0.211 (0.166, 0.263) | 1.77 |
| | Baykan | 2012 | 0.325 (0.234, 0.432) | 1.64 |
| | Chan | 2011 | 0.156 (0.105, 0.225) | 1.72 |
| | Azegbeobor | 2015 | 0.175 (0.124, 0.241) | 1.73 |
| | Sumanathissa | 2011 | 0.279 (0.211, 0.358) | 1.72 |
| | Preljevic | 2013 | 0.220 (0.153, 0.307) | 1.68 |
| | Cruz | 2010 | 0.357 (0.255, 0.474) | 1.61 |
| | Koo | 2003 | 0.548 (0.425, 0.666) | 1.58 |
| | El Filali | 2017 | 0.340 (0.256, 0.436) | 1.68 |
| | Watnick | 2005 | 0.258 (0.166, 0.379) | 1.58 |
| | Kalender | 2006 | 0.241 (0.178, 0.318) | 1.72 |
| | Hinrichsen | 1989 | 0.242 (0.175, 0.324) | 1.70 |
| | Loosman | 2010 | 0.339 (0.233, 0.463) | 1.58 |
| | de Alencar | 2019 | 0.225 (0.170, 0.293) | 1.74 |
| | Aghanwa | 1997 | 0.350 (0.181, 0.567) | 1.23 |
| | Gadia | 2020 | 0.660 (0.563, 0.745) | 1.67 |
| | Choi | 2012 | 0.506 (0.400, 0.612) | 1.64 |
| | Niharika | 2021 | 0.452 (0.334, 0.575) | 1.58 |
| | Hamody | 2013 | 0.800 (0.696, 0.875) | 1.62 |
| | Martens | 2018 | 0.054 (0.046, 0.062) | 1.84 |
| | Wang | 2014 | 0.239 (0.184, 0.305) | 1.75 |
| | Rodriguez-Angarita | 2016 | 0.080 (0.052, 0.120) | 1.77 |
| | Tuna | 2021 | 0.393 (0.276, 0.524) | 1.56 |
| | Agrawal | 2019 | 0.510 (0.413, 0.606) | 1.67 |
| | Kokoszka | 2016 | 0.785 (0.698, 0.852) | 1.68 |
| | Macaron | 2014 | 0.490 (0.359, 0.623) | 1.54 |
| | Kalender | 2007 | 0.262 (0.153, 0.411) | 1.49 |
| | **Subgroup, DL ($I^2$ = 97.4%, p < 0.001)** | | **0.280 (0.242, 0.319)** | **97.31** |
| Paediatric | Bakr | 2007 | 0.105 (0.042, 0.241) | 1.46 |
| | Berney-Martinet | 2018 | 0.250 (0.112, 0.469) | 1.23 |
| | **Subgroup, DL ($I^2$ = 48.6%, p = 0.163)** | | **0.159 (0.042, 0.323)** | **2.69** |
| **Heterogeneity between groups: p = 0.167** | | | | |
| **Overall, DL ($I^2$ = 97.3%, p < 0.001)** | | | **0.276 (0.239, 0.315)** | **100.00** |

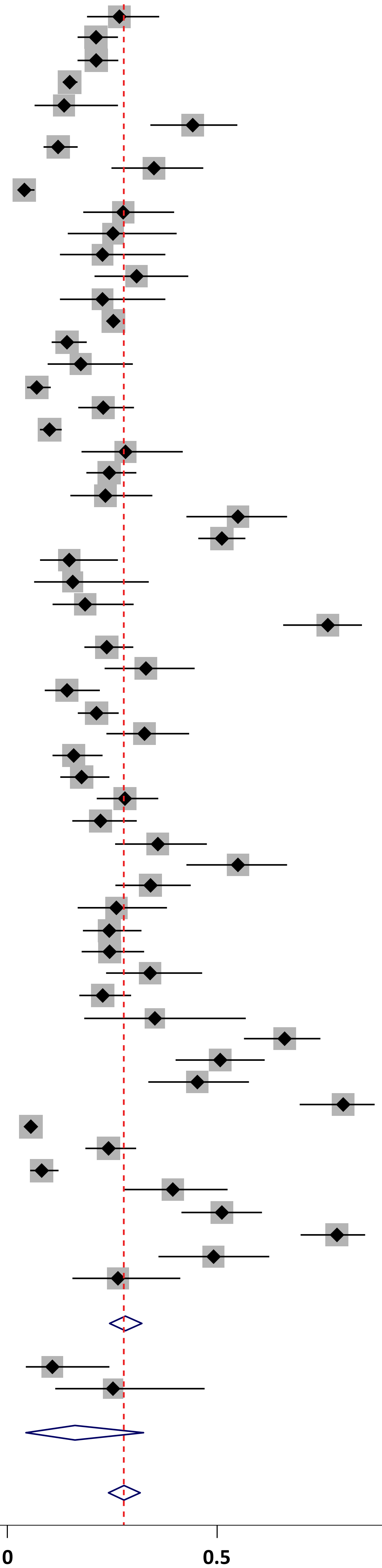


**NOTE**: Weights and between-subgroup heterogeneity test are from random-effects model

**Supplementary Fig. 4:** Forest plot comparing the pooled prevalence of depression in HD versus PD dialysis patients

| Group | Author | Year | Proportion (95% CI) | Weight(%) |
|---|---|---|---|---|
| HD | Hedayati | 2006 | 0.265 (0.188, 0.360) | 2.32 |
| HD | Hedayati | 2005 | 0.147 (0.130, 0.165) | 2.46 |
| HD | Bautovich | 2018 | 0.133 (0.063, 0.262) | 2.17 |
| HD | Gupta | 2018 | 0.440 (0.339, 0.547) | 2.30 |
| HD | Jose | 2006 | 0.119 (0.084, 0.165) | 2.41 |
| HD | Drayer | 2006 | 0.274 (0.179, 0.396) | 2.24 |
| HD | Cilan | 2012 | 0.225 (0.123, 0.375) | 2.14 |
| HD | Balogun | 2011 | 0.306 (0.206, 0.430) | 2.24 |
| HD | Chilcot | 2008 | 0.225 (0.123, 0.375) | 2.14 |
| HD | Loureiro | 2018 | 0.140 (0.103, 0.187) | 2.42 |
| HD | Reckert | 2013 | 0.173 (0.094, 0.297) | 2.21 |
| HD | Fukunishi | 2002 | 0.098 (0.075, 0.127) | 2.44 |
| HD | Soykan | 2004 | 0.280 (0.175, 0.417) | 2.20 |
| HD | Yeh | 2014 | 0.241 (0.186, 0.306) | 2.40 |
| HD | Martiny | 2011 | 0.232 (0.148, 0.344) | 2.27 |
| HD | Koo | 2005 | 0.548 (0.425, 0.666) | 2.24 |
| HD | Saritha | 2021 | 0.510 (0.454, 0.566) | 2.42 |
| HD | Dimaano | 2021 | 0.145 (0.076, 0.262) | 2.22 |
| HD | Alsuwaida | 2006 | 0.154 (0.062, 0.335) | 1.99 |
| HD | Chandra | 2011 | 0.183 (0.106, 0.299) | 2.24 |
| HD | Donia | 2015 | 0.763 (0.656, 0.845) | 2.28 |
| HD | Chen | 2010 | 0.235 (0.182, 0.298) | 2.40 |
| HD | Cukor | 2008 | 0.329 (0.230, 0.445) | 2.27 |
| HD | Huang | 2007 | 0.140 (0.087, 0.218) | 2.34 |
| HD | Azegbeobor | 2015 | 0.175 (0.124, 0.241) | 2.38 |
| HD | Bakr | 2007 | 0.105 (0.042, 0.241) | 2.12 |
| HD | Cruz | 2010 | 0.357 (0.255, 0.474) | 2.27 |
| HD | Koo | 2003 | 0.548 (0.425, 0.666) | 2.24 |
| HD | El Filali | 2017 | 0.340 (0.256, 0.436) | 2.33 |
| HD | Hinrichsen | 1989 | 0.242 (0.175, 0.324) | 2.35 |
| HD | de Alencar | 2019 | 0.225 (0.170, 0.293) | 2.39 |
| HD | Aghanwa | 1997 | 0.350 (0.181, 0.567) | 1.88 |
| HD | Gadia | 2020 | 0.660 (0.563, 0.745) | 2.33 |
| HD | Choi | 2012 | 0.506 (0.400, 0.612) | 2.29 |
| HD | Niharika | 2021 | 0.452 (0.334, 0.575) | 2.24 |
| HD | Hamody | 2013 | 0.800 (0.696, 0.875) | 2.28 |
| HD | Wang | 2014 | 0.239 (0.184, 0.305) | 2.39 |
| HD | Tuna | 2021 | 0.393 (0.276, 0.524) | 2.22 |
| HD | Agrawal | 2019 | 0.510 (0.413, 0.606) | 2.33 |
| HD | Kokoszka | 2016 | 0.785 (0.698, 0.852) | 2.34 |
| HD | Macaron | 2014 | 0.490 (0.359, 0.623) | 2.20 |
| HD | **Subgroup, DL ($I^2$ = 95.6%, p < 0.001)** | | **0.319 (0.260, 0.381)** | **93.35** |
| PD | Cilan | 2013 | 0.250 (0.142, 0.402) | 2.14 |
| PD | Chan | 2011 | 0.156 (0.105, 0.225) | 2.37 |
| PD | Kalender | 2007 | 0.262 (0.153, 0.411) | 2.15 |
| PD | **Subgroup, DL ($I^2$ = 41.7%, p = 0.180)** | | **0.204 (0.131, 0.287)** | **6.65** |
| | **Heterogeneity between groups: p = 0.028** | | | |
| | **Overall, DL ($I^2$ = 95.3%, p < 0.001)** | | **0.312 (0.256, 0.370)** | **100.00** |

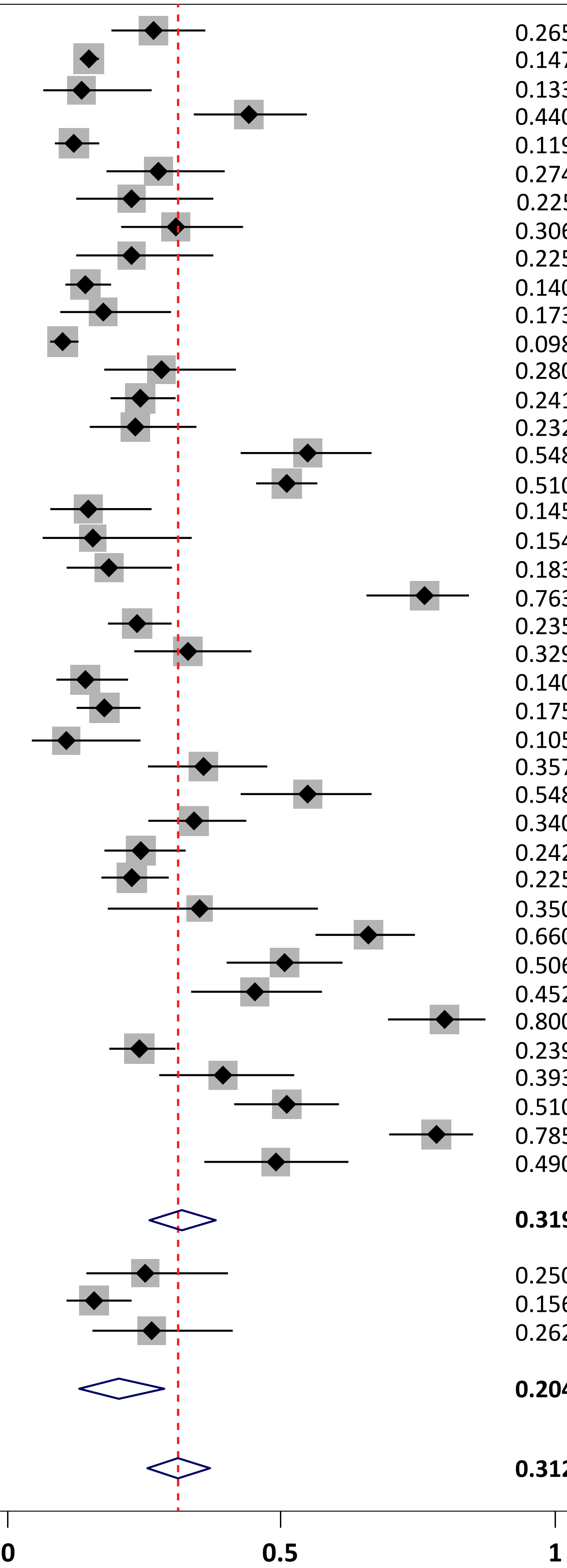


**NOTE**: Weights and between-subgroup heterogeneity test are from random-effects model

**HD** = Hemodialysis; **PD** = Peritoneal Dialysis

### PRISMA 2009 Checklist

| Section/topic | # | Checklist item | Reported on page # |
|---|---|---|---|
| **TITLE** | | | |
| Title | 1 | Identify the report as a systematic review, meta-analysis, or both. | 1 |
| **ABSTRACT** | | | |
| Structured summary | 2 | Provide a structured summary including, as applicable: background; objectives; data sources; study eligibility criteria, participants, and interventions; study appraisal and synthesis methods; results; limitations; conclusions and implications of key findings; systematic review registration number. | 2 |
| **INTRODUCTION** | | | |
| Rationale | 3 | Describe the rationale for the review in the context of what is already known. | 3 |
| Objectives | 4 | Provide an explicit statement of questions being addressed with reference to participants, interventions, comparisons, outcomes, and study design (PICOS). | 3 |
| **METHODS** | | | |
| Protocol and registration | 5 | Indicate if a review protocol exists, if and where it can be accessed (e.g., Web address), and, if available, provide registration information including registration number. | 3 |
| Eligibility criteria | 6 | Specify study characteristics (e.g., PICOS, length of follow-up) and report characteristics (e.g., years considered, language, publication status) used as criteria for eligibility, giving rationale. | 3-4 |
| Information sources | 7 | Describe all information sources (e.g., databases with dates of coverage, contact with study authors to identify additional studies) in the search and date last searched. | 3-4 |
| Search | 8 | Present full electronic search strategy for at least one database, including any limits used, such that it could be repeated. | Supplementary Tables 2A & B |
| Study selection | 9 | State the process for selecting studies (i.e., screening, eligibility, included in systematic review, and, if applicable, included in the meta-analysis). | 4 |
| Data collection process | 10 | Describe method of data extraction from reports (e.g., piloted forms, independently, in duplicate) and any processes for obtaining and confirming data from investigators. | 4 |
| Data items | 11 | List and define all variables for which data were sought (e.g., PICOS, funding sources) and any assumptions and simplifications made. | 4 |
| Risk of bias in individual studies | 12 | Describe methods used for assessing risk of bias of individual studies (including specification of whether this was done at the study or outcome level), and how this information is to be used in any data synthesis. | 4 |
| Summary measures | 13 | State the principal summary measures (e.g., risk ratio, difference in means). | 4 |
| Synthesis of results | 14 | Describe the methods of handling data and combining results of studies, if done, including measures of consistency (e.g., $I^2$) for each meta-analysis. | 4 |

### PRISMA 2009 Checklist



| Section/topic | # | Checklist item | Reported on page # |
|---|---|---|---|
| Risk of bias across studies | 15 | Specify any assessment of risk of bias that may affect the cumulative evidence (e.g., publication bias, selective reporting within studies). | 4 |
| Additional analyses | 16 | Describe methods of additional analyses (e.g., sensitivity or subgroup analyses, meta-regression), if done, indicating which were pre-specified. | 4 |
| **RESULTS** | | | |
| Study selection | 17 | Give numbers of studies screened, assessed for eligibility, and included in the review, with reasons for exclusions at each stage, ideally with a flow diagram. | 4-5 |
| Study characteristics | 18 | For each study, present characteristics for which data were extracted (e.g., study size, PICOS, follow-up period) and provide the citations. | 4-5 |
| Risk of bias within studies | 19 | Present data on risk of bias of each study and, if available, any outcome level assessment (see item 12). | 4-5 |
| Results of individual studies | 20 | For all outcomes considered (benefits or harms), present, for each study: (a) simple summary data for each intervention group (b) effect estimates and confidence intervals, ideally with a forest plot. | Tables 1-4 |
| Synthesis of results | 21 | Present results of each meta-analysis done, including confidence intervals and measures of consistency. | 4-5 |
| Risk of bias across studies | 22 | Present results of any assessment of risk of bias across studies (see Item 15). | 4-5 |
| Additional analysis | 23 | Give results of additional analyses, if done (e.g., sensitivity or subgroup analyses, meta-regression [see Item 16]). | 5 |
| **DISCUSSION** | | | |
| Summary of evidence | 24 | Summarize the main findings including the strength of evidence for each main outcome; consider their relevance to key groups (e.g., healthcare providers, users, and policy makers). | 7-8 |
| Limitations | 25 | Discuss limitations at study and outcome level (e.g., risk of bias), and at review-level (e.g., incomplete retrieval of identified research, reporting bias). | 7-8 |
| Conclusions | 26 | Provide a general interpretation of the results in the context of other evidence, and implications for future research. | 7-8 |
| **FUNDING** | | | |
| Funding | 27 | Describe sources of funding for the systematic review and other support (e.g., supply of data); role of funders for the systematic review. | 8 |

*From:* Moher D, Liberati A, Tetzlaff J, Altman DG, The PRISMA Group (2009). Preferred Reporting Items for Systematic Reviews and Meta-Analyses: The PRISMA Statement. PLoS Med 6(7): e1000097. doi:10.1371/journal.pmed1000097

For more information, visit: [www.prisma-statement.org](http://www.prisma-statement.org).

**Supplementary Table 2A: Search Strategy on PubMed**

| Search Number | Query | Result |
|---|---|---|
| **4** | ((#1) AND (#2)) AND (#3) | *3,768* |
| **3** | (((((((((Chronic Kidney Disease) OR (CKD)) OR (chronic renal disease)) OR (chronic kidney failure)) OR (end-stage renal disease)) OR (end-stage kidney disease)) OR (chronic renal insufficiency)) OR (chronic renal failure)) OR (end-stage kidney failure)) OR (chronic renal impairment)) OR (kidney failure) | *340,053* |
| **2** | ((((((((depression) OR (low mood)) OR (dysthymia)) OR (persistent depressive disorder)) OR (major depressive disorder)) OR (minor depressive disorder)) OR (anhedonia)) OR (loss of interest in pleasurable activities)) OR (low energy) | *884,383* |
| **1** | ((((((prevalence) OR (frequency)) OR (incidence)) OR (occurrence)) OR (burden)) OR (preponderance)) OR (rate) | *7,196,316* |

**Supplementary Table 2B: Search Strategy on Embase**

| Search Number | Query | Result |
|---|---|---|
| **1** | Prevalence OR frequency OR incidence OR occurrence OR burden OR preponderance OR rate | *7,617,790* |
| **2** | Depression OR low mood OR dysthymia OR persistent depressive disorder OR major depressive disorder OR minor depressive disorder OR anhedonia OR loss of interest in pleasurable activities OR low energy | *854,699* |
| **3** | Chronic Kidney Disease OR CKD OR chronic renal disease OR chronic kidney failure OR end-stage renal disease OR end-stage kidney disease OR chronic renal insufficiency OR chronic renal failure OR (end-stage kidney failure OR chronic renal impairment OR kidney failure | *505,903* |
| **4** | 1 AND 2 AND 3 | *5,406* |
